# COMPARISON OF PERFORMANCE CHARACTERISTICS BETWEEN LATERAL FLOW, ELISA AND ELECTROCHEMILUMINESCENCE IMMUNOASSAYS FOR THE DETECTION OF SARS-COV-2 ANTIBODIES AMONG HEALTHCARE WORKERS

**DOI:** 10.1101/2021.04.29.21256260

**Authors:** M R Shincy, Vandana Govindan, H H Sudhakar, V T Venkatesha, K Padmapriya, K L Ravikumar

## Abstract

**Background:** Medical professionals and researchers have been urging the need for wide and rapid testing of citizens in order to plan measures that can contain the spread of the virus. Antibody tests play an important role throughout the patient care pathway and are vital for the management and surveillance of the virus. Although RT-PCR is considered as the gold standard, serological tests based on antibodies are helpful for on-time detection. We performed one to one assessment of point-of-care lateral flow assay (POCTs), enzyme immunoassay (EIAs), electrochemiluminescence immunoassay (CLIA), to detect severe acute respiratory syndrome coronavirus 2 (SARS-CoV-2) IgG antibody.

**Materials and Methods:** 611 healthcare workers were recruited between November and December 2020 at Central Research Laboratory, KIMS. Collected serum samples were analysed according to manufacturer’s protocol. The Standard Q IgG/IgM combo assay, Anti-SARS CoV-2 Human IgG ELISA, and the Elecsys^®^ to measure the IgG titer of severe acute respiratory syndrome coronavirus 2 (SARS-CoV-2).

**Results:** The kits displayed a sensitivity of 61.2%,79.5%, 91.8% and specificity of 61.7%,64.1%,80.2% for the Standard Q IgG/IgM combo assay, Anti-SARS CoV-2 Human IgG ELISA, and the Elecsys^®^ in order.

**Conclusion:** Our results indicate high sensitivity and specificity for the Elecsys^®^ assay compared to Anti-SARS CoV-2 Human IgG ELISA, the Standard Q IgG/IgM combo assay.

## 1. INTRODUCTION

The severe acute respiratory syndrome coronavirus 2 (SARS-CoV-2) first reported in late 2019 has caused a large global outbreak and is a major public health issue ^(1)^. Early and accurate diagnosis of SARS-CoV-2 infection is essential for prevention and pandemic containment. The heterogeneity of the clinical presentation, from asymptomatic individuals to severe cases, and the diversity of clinical manifestations of COVID-19, emphasize the need for tests with good sensitivity and specificity ^(2)^. Real-Time Reverse Transcriptase Polymerase Chain Reaction (RT-PCR) assays detecting the presence of viral genetic material are presently gold-standard of diagnosis. However, several cases of false-negative results have been described due to low viral load, complex techniques, inappropriate sample collection ^(3)^. The outcome can be dramatic with infectious patients spreading the viruses hampering attempts by public health to contain viral circulation ^(4)^.

Unlike molecular testing, detection of an immune response to the virus is an indirect marker of infection. A wide range of serology immunoassays with different SARS-CoV-2 antigen recognition and antibody specificity has been developed to compliment RT-PCR assays ^(5,6)^. Accurate and high throughput serological assays facilitate large-scale studies and improve the understanding of the true proportion of the population that has recovered from COVID-19. The testing for the presence of antibodies on a widespread scale could help evidence-based decision-making on an individual and social scale, minimizing the economic impact of the virus. Several different antibody tests have been developed, which differ depending on the targeted viral antigen (for example, nucleoprotein or spike protein), biomarkers tested IgM or IgG antibodies ^(7,8)^.

After infection with SARS-CoV-2, the immune system identifies and produces an immune response by generating unique antibodies. The immunoglobulin M (IgM) antibody, produced in the early period after the infection, can indicate the current or the recent infection. Immunoglobulin G (IgG) antibody indicates that the disease is in the middle to late stage or presence of past infection. Detection of IgG antibodies against SARS-CoV-2 has a significant role as it is long-lasting and correlated with viral neutralizing activity, which is important for recovery ^(9,10,)^. Test results indicate that IgG produced against SARS-CoV-2 antigens is detectable in immunocompetent patients at least 8 days after onset of symptoms, with more than 90 percent of subjects seropositive after 14 days of illness. Some individuals can take longer to seroconvert depending on their immune status or may never be seroconverted if substantially immunosuppressed ^(11,12)^. Initial studies indicate a relatively high specificity (95%) for IgG-based serological assays against circulating coronaviruses ^(13,14,15)^.

Emerging SARS-CoV-2 Viral disease has a huge influence on health staff. The risk profile for SARS-CoV-2 exposure and infection among health workers varies considerably ^(16)^. Health care workers are likely to be in touch with patients and co-workers who have atypical, little or no signs while also being extremely infectious ^(17)^. They are thus a high-risk group, and studies linked to their seroprevalence, asymptomatic cases, seroconversion, and transmission of the disease are crucial. SARS-CoV-2 Serological assays are variable, with varying formats. Many laboratories have minimum information about performance limitations for choosing the assay due to limited Peer-reviewed literature on serological testing currently available. Independent validation is also needed to ensure the assays are in line with expected analytical and clinical performance specifications. The best use of SARS-CoV-2 serological testing remains an open issue.

The present study aimed to compare the performance of lateral flow immunoassays [LFIAs] Standard Q IgG/IgM combo assay, enzyme-linked immunosorbent assays [ELISAs] Anti-SARS CoV-2 Human IgG ELISA, and chemiluminescent immunoassays [CLIA] Elecsys ^®^ anti-SARS-CoV-2 immunoassays in healthcare workers.

## 2. MATERIALS AND METHODS

### 2.1 Materials

This study was conducted at Central Research Laboratory, Kempegowda Institute of Medical Science Hospital& Research Centre (CRL, KIMSH&RC) and the protocol has the approval of institutional ethics committee KIMS/IEC/A044/M/2020. A total of 611 blood samples were collected from healthcare workers including Physicians, Nurses, Laboratory Technologists, Pharmacists, Administrative, and Housekeeping staff working in different sections of KIMSH&RC (Table 1). Written informed consent was taken from the participants. In a questionnaire demographic information, hand hygiene habits, use of PPEs were documented. Information about participant exposure to covid patients, duration of exposure, history of covid infection, RTPCR results, and Co-morbidities were recorded. 2 ml of venous blood collected from enrolled participants was centrifuged and separated serum was stored at -20^□^C. In our study, all the assays were evaluated with the same panel of samples under similar conditions to analyze their performance. In the study, I) Standard Q covid-19 IgG/IgM combo test II) Anti-SARS CoV-2 Human IgG ELISA III) Elecsys ^®^ tests were evaluated for their sensitivity and specificity.

**Table 1:** Participants details

| **Participants** | **N (611)** |
| --- | --- |
| Doctors | 261 |
| Nurses | 129 |
| Lab Personnel | 87 |
| Paramedics | 76 |
| Ancillary staff | 58 |

### 2.2 Overview of Immunoassay methods

#### I) Lateral flow immunoassay

The Standard Q Covid -19 IgG/IgM Combo test is a rapid chromatographic immunoassay for the qualitative detection of antibodies manufactured by SD biosensor. During the test, SARS-CoV-2 antibodies in the specimen interact with the recombinant COVID-19 nucleocapsid protein conjugated with the colloidal gold particle that makes the antigen-antibody gold particle complex. This complex migrates on the membrane via capillary action until the “M” and “G” test line, where it will be captured by the monoclonal anti-human IgM antibody or anti-human IgG antibody. Goat polyclonal anti-mouse IgG antibody is coated on the control line region and monoclonal anti - human IgG antibody and monoclonal anti-human IgM antibody is coated on the “G” and “M” test line. If no antibodies are present in the sample, then no color appears in the test line.

#### II) Enzyme Linked Immunosorbent Assay

Covid Kavach IgG ELISA kit developed by the Indian Council of Medical Research’s National Institute of Virology is licensed for manufactured by Meril Diagnostics. It qualitatively detects IgG antibodies that bind to the SARS CoV-2 virus whole-cell antigen coated onto the microtiter plate. The kit suggests an interpretation of the results based on the OD value. If the OD value is >Cut off it is considered as Positive and the OD value is <Cut off it is Negative ^(18)^.

#### III) Electrochemiluminescence immunoassay

Elecsys® Anti-SARSCoV-2 electrochemiluminescence immunoassay qualitatively detects SARS-CoV-2 antibodies. We measured SARS-CoV-2 antibodies automated on the Cobas e411 analyzer (Roche Diagnostics). This assay is a modified double-antigen sandwich immunoassay using recombinant nucleocapsid protein (N), which detects the antibody. Results are reported as positive if the OD is >1 and Negative if the OD is <1. ^(19)^.

The three assays are compared for the analyte, antigen, and the methods (Table 2). These tests were performed as per the manufacturer’s instructions.

**Table 2:** Characteristics of Commercial Anti-SARS-Cov-2 Assay Used in This Comparative Study

| Manufacturers | Assay | Analyte | Capture Antigen | Method | Analyzer | Cut off values |  |  |
| --- | --- | --- | --- | --- | --- | --- | --- | --- |
|  |  |  |  |  |  | Negative | Indeterminate | Positive |
| SD Biosensor | Standard Q IgG/IgM combo assay | IgG & IgM | Nucleocapsid | LFA | NA | Only band in the control (C) line | No band in Control (C) line | coloured bands (G line) |
| Meril | Anti-SARS CoV-2 Human IgG | IgG | Whole-cell antigen | ELISA | Alere easy reader | < Cutoff | NA | >Cutoff |
| Roche | Elecsys<sup>®</sup> anti-SARS-CoV-2 | Total immunoglobulin | Nucleocapsid | CLIA | Cobas e411 | <1 | NA | ≥1 |

## 3. RESULTS

### 3.1 Population demographics

611 sera were included in this study, 538 were collected from patients with a negative SARS-CoV-2 PCR result and 73 from patients with a positive SARS-CoV-2 PCR result in serum samples. The mean age of patients was 36.5 (range 17 to 68 years) (Table 3), with a majority being Female (65.1%) (Fig 1). The most common comorbidities in the cohort were Obesity (71.3%), hypertension (44.4%), Diabetes (35.4%), and asthma (20%) (Fig2).

**Table 3:** Age distribution among the participants

| **Age Group** | **N (611)** |
| --- | --- |
| 10-20 | 01 |
| 20-29 | 189 |
| 30-39 | 140 |
| 40-49 | 125 |
| 50-59 | 110 |
| 60-69 | 46 |

**Fig 1:**
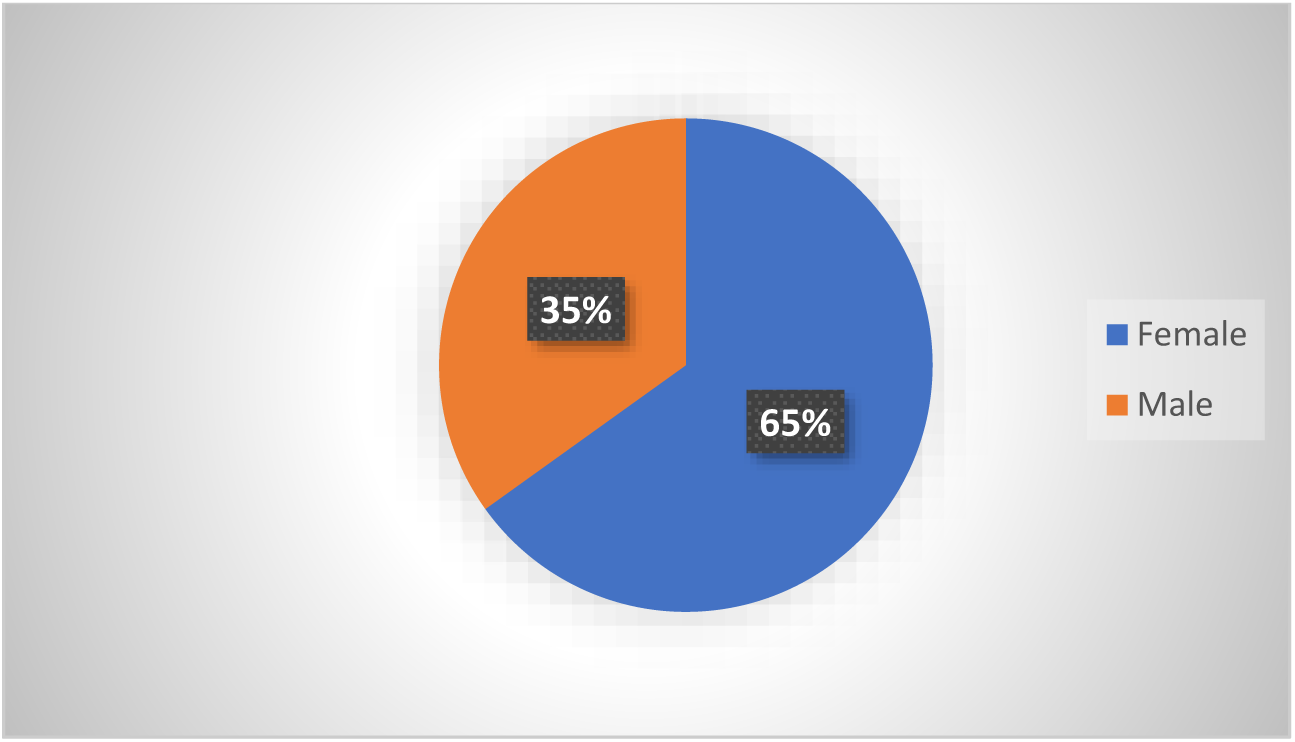
Male-Female ratio

**FIG 2;.**
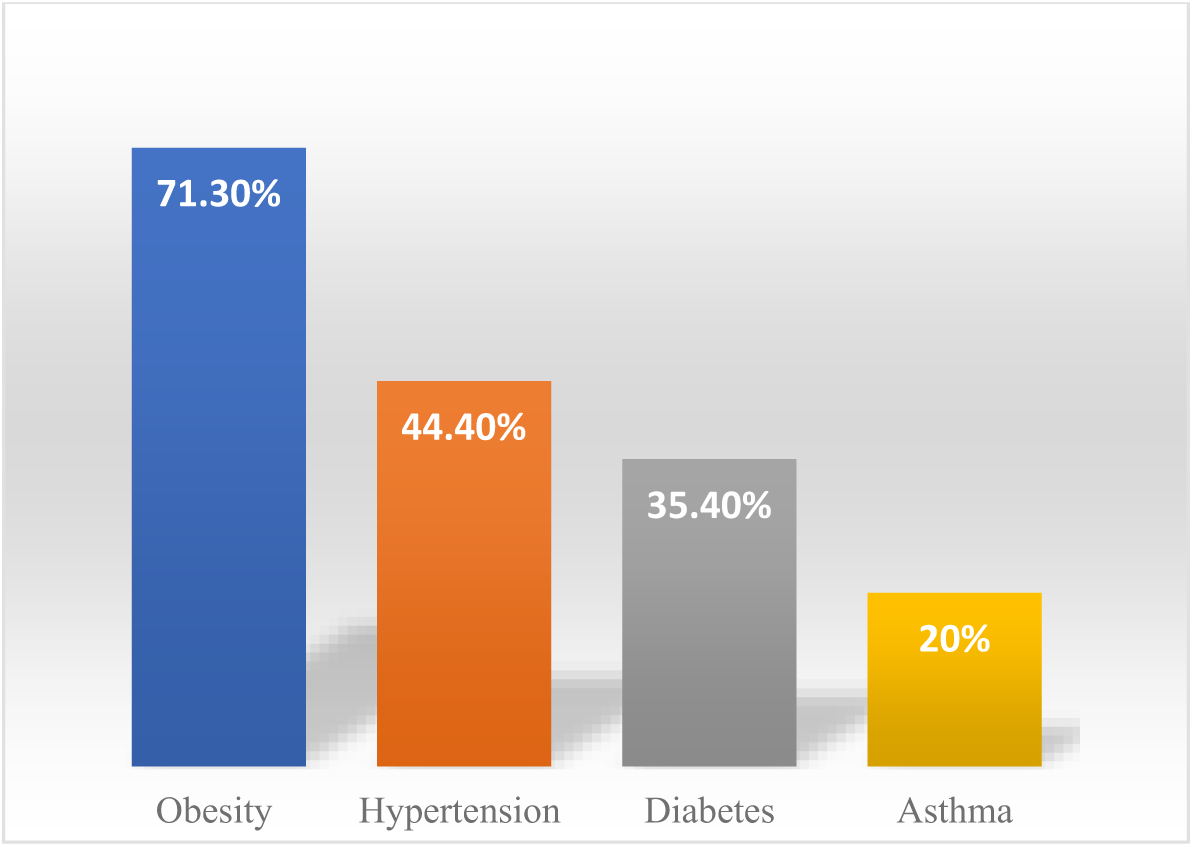
Percentage of Co-morbidities

### 3.2 Determination of sensitivity and specificity of the assays

Each of the 611 serum samples was tested in parallel with three assays for the presence of SARS-CoV-2 IgG antibodies as recommended by the manufacturers. The Standard Q covid-19 IgG/IgM combo assay revealed 79 positive test results and 532 negative results Anti-SARS CoV-2 Human IgG ELISA showed 160 SARS CoV-2 IgG Positive 451 Negatives. With Elecsys^®^ assay, 107 sera had a positive result, and 504 had a negative result.

The sensitivity was 61.2%, 79.5%, and 91.8% for Standard Q covid-19 IgG/IgM combo, Anti-SARS CoV-2 Human IgG ELISA, and Elecsys^®^ respectively. 61.7%,64.1%, and 80.2% were the specificity of the assays in order (Table 4).

**Table 4:** Percentage of Sensitivity and Specificity

| ASSAY | Sensitivity | Specificity |
| --- | --- | --- |
| Lateral flow immunoassays | 61.2% | 61.7% |
| ELISA | 79.5% | 64.1% |
| Electrochemiluminescence | 91.8% | 80.2% |

## 4. DISCUSSION

As the spread of COVID-19 continues, precise antibody tests are essential for public health interventions to monitor and assess the level of immunity within a given population. SARS-CoV-2 specific antibodies appear a week after onset of symptoms, supplementing the diagnostic repertoire in identifying patients with past infection. Knowing the antibody levels helps understand the prevalence of infection in the population, the timeline of antibody development in different patient populations, the longevity of the antibody response, and the course of the pandemic ^(20)^. Besides, it identifies presumably immune health care workers who can work with the patients, plasma donors for therapeutic transfusion, and detection of patients presenting in the later stage of disease with viral clearance preceding the disappearance of symptoms ^(21, 22)^. This study was executed during November-December 2020 by Central Research Laboratory, KIMS to compare the performance of commercially available kits for the detection of SARS-CoV-2 IgG antibodies.

Serological tests detect the antibodies produced by the immune response to infection like COVID-19. The lag phase of primary response to the antigenic stimulus like virus varies from 4 to 10 days with IgM predominance followed by IgG. The precision of antibody tests in COVID-19 has been a topic of debate. Since the magnitude of antibody response depends on the severity of infection; patients with asymptomatic infections/mild infections may not build up a measurable antibody response ^(23)^. This limits the test’s effectiveness in diagnosing the COVID-19 during the early stage of symptoms. However serological test plays an important role to identify and follow individuals who have developed an immune response against infection and to vaccines. In addition, test result provides information about the severity of infection blood donors who can donate convalescent anti covid plasma and herd immunity.

Numerous immunoassays for the detection of antibodies to SARS-CoV-2 are currently in development or commercially available. Broadly they belong to three groups a) Point-of-care, Lateral flow assays (LFIA) b) Enzyme-Linked Immunosorbent Assays (ELISA) c) Chemiluminescence assays (CLIA) However, not all antibody tests are marketed to the public have been evaluated. Published studies vary dramatically in their sensitivity and specificity of these assays and many lack information regarding target antigen and negative control groups ^(24,25)^. It is important to understand their performance characteristics, limitations, and whether these antibodies convey immunity that would prevent or reduce the severity of re-infection as well as the duration for which immunity lasts ^(26,27)^. There is an urgent need to validate these assays taking into account variables that will affect the sensitivity and specificity of the different immunoassays. The major issue faced by the commercially available assays in the market is the very little correlation in their sensitivity and specificity. The present study was designed to evaluate the sensitivity and specificity of three different methodologies for COVID 19 antibody testing with kits available in the open market.

Lateral flow immunoassays have the advantage of decreased technical requirement, short turnaround time, affordability, lower sampling, and specimen preparation risk. Lateral flow immunoassays have been developed to detect anti covid IgM and IgG antibodies separately or simultaneously ^(28)^. With a large number of commercially available devices from different companies, variable sensitivity and specificity are reported in the publications ^(29,30,31)^. Serum sample set of 611 individuals obtained from 73 positive patients defined by positive RT-PCR and covid 19 symptoms and 538 negatives were evaluated with LFIA. The assay sensitivity was 61.2% and the specificity was 61.7%. Our study reports the lowest sensitivity of LFIAs among the three tests which is less than reported by the manufacturer. A similar low sensitivity rate for the LFIAs method is reported in a recent meta-analysis on antibody tests ^(32)^. The ELISA technique offers the advantage of determining the antibody titers and isotype detection ^(33)^. However, as the technique is labour-intensive with the need for equipment is unsuitable for POCT. The assay sensitivity was 79.5% with a specificity of 64.1% in our samples. The CLIA assay is traditionally considered a very sensitive method with the capability to detect low levels of antibodies ^(34)^. These assays are automated and allow for a high throughput of samples. CLIA test results were 91.2% sensitivity and 81.2%specificity. In the study best correlation was found between CLIA and ELISA.

The Limitations of the study were the moderate numbers of positive samples, evaluating with genomic RT-PCR and failure to take into consideration time of sample collection from the initial symptoms and RT-PCR testing as these antibody tests typically become useful two weeks after initial symptoms.

## 5. CONCLUSION

The strength of this study is the side-by-side evaluation of three assays with a large number of negative samples to give reliable and comparable specificity data. Among the three tests, the Elecsys^®^ assay exhibited high specificity and sensitivity for identifying IgG antibodies. LFIA is an appealing platform with a relatively low cost per test and convenience. Understanding the strength and pitfall of these assays in different clinical and research scenarios are critically needed for their utilization of serological testing.

## Data Availability

The data that support the findings of this study are available from the corresponding author, [Shincy M R], upon reasonable request

## Compliance with ethical standards

### Ethics approval

The study was conducted according to the ethical requirements established by the Declaration of Helsinki. The Ethics Committee of Kempegowda Institute of Medical Science approved the study.

## References

1. Wei Feng, Wei Zong, Feng Wang & Shaoqing Ju. Severe acute respiratory syndrome coronavirus 2 (SARS-CoV-2): a review. Molecular Cancer 2020: 19 –100.

2. Buddhisha Udugama, Pranav Kadhiresan, Hannah N. Kozlowski,Ayden Malekjahani,Matthew Osborne and Vanessa Y. C. Li et.al. Diagnosing COVID-19: The Disease and Tools for Detection. ACS Nano ;2020;14(4):3822 –3835.

3. Junxiong Pang, Min Xian Wang, Ian Yi Han Ang, Sharon Hui Xuan Tan, Ruth Frances Lewis and Jacinta I-Pei Chen et.al. Potential Rapid Diagnostics, Vaccine and Therapeutics for 2019 Novel Coronavirus (2019-nCoV): A Systematic Review. J. Clin. Med 2020; 9:623.

4. Shuo Su, Gary Wong, Weifeng Shi, Jun Liu, Alexander C.K. Lai, and Jiyong Zhou et.al. Epidemiology, Genetic Recombination, and Pathogenesis of Coronaviruses. Trends in Microbiology 2016 Jun;24(6):490 –502.

5. Yujiao Jina, Miaochan Wanga, Zhongbao Zuoa, Chaoming Fanb, Fei Y ec and Zhaobin Cai et.al. Diagnostic value and dynamic variance of serum antibody in coronavirus disease 2019. International Journal of Infectious Diseases 2020; 94:49 –52.

6. Sarfaraz Ahmad Ejazi, Sneha Ghosh and Nahid Ali. Antibody detection assays for COVID‐19 diagnosis: an early overview. Immunology & Cell Biology 2021 Jan;99(1):21–33

7. Jacofsky, D., Jacofsky, E. M., & Jacofsky, M. Understanding Antibody Testing for COVID-19. The Journal of Arthroplasty ; 35(7S), S74–S81 (2020)

8. Arturo Casadevall and Liise-anne Pirofski The convalescent sera option for containing COVID-19. Clin Invest. 2020;130(4):1545 –1548.

9. Li-Xiang Wu, Hui Wang, Dan Gou, Gang Fu, Jing Wang and Bian-Qin Guo. Clinical significance of the serum IgM and IgG to SARS‐CoV‐2 in coronavirus disease‐2019. Clin Lab Anal. 2020;00: e23649.

10. Jonathan Kopel, Hemant Goyal and Abhilash Perisetti. Antibody tests for COVID-19. Baylor University Medical Center Proceedings 2020;0(0):1 –10.

11. Quan-xin Long a, Hai-jun Deng, Juan Chen, Jie-li Hu a, Bei-zhong Liu c, and Pu Liao et.al Antibody responses to SARS-CoV-2 in COVID-19 patients: the perspective application of serological tests in clinical practice. Clin Infect Dis. 2020 Nov 19;71(16):2027–2034.

12. Juanjuan Zhao, Quan Yuan, Haiyan Wang,Wei Liu, Xuejiao Liao and Yingying Su. Antibody responses to SARS-CoV-2 in patients of novel coronavirus disease 2019. Clin Infect Dis 2020 Nov 19;71(16):2027 –2034.

13. Alexander Krüttgen, Christian G Cornelissen, Michael Dreher, Mathias Hornef, Matthias Imöhl and Michael Kleines. Comparison of four new commercial serologic assays for determination of SARS-CoV-2 IgG. Journal of Clinical Virology 2020 ;128: 104394.

14. Carmen L. Charlton, Jamil N. Kanji, Kam Johal and Ashley Bailey. Evaluation of Six Commercial Mid-to High-Volume Antibody and Six Point-of-Care Lateral Flow Assays for Detection of SARS-CoV-2 Antibodies. Journal of Clinical Microbiology 2020.

15. Etienne Brochot, Baptiste Demey, Lynda Handala,Catherine François, Gilles Duverlie, and Sandrine Castelain.Comparison of different serological assays for SARS-CoV-2 in real life. Journal of Clinical Virology 2020; 130: 104569.

16. Julia A Bielicki, Xavier Duval, Nina Gobat, Herman Goossens, Marion Koopmans and Evelina Tacconelli.Monitoring approaches for health-care workers during the COVID-19 pandemic. Personal view volume 20.

17. Ann-Sofie Rudberg, Sebastian Havervall, Anna Månberg, August Jernbom Falk, Katherina Aguilera, and Henry Ng et.al SARS-CoV-2 exposure, symptoms and seroprevalence in healthcare workers in Sweden. Nature Communications 2020; 11:5064.

18. ICMR-NIV anti-SARS-COV-2 human IgG elisa covid kavach-merilisa: ckmeli-01.

19. Elecsys® Anti-SARS-CoV-2:09203095190

20. Katharina Röltgen, Abigail E. Powell, Oliver F. Wirz, Bryan A. Stevens, Catherine A. Hogan,Javaria Najeeb,Molly Hunter et.al. Defining the features and duration of antibody responses to SARS-CoV-2 infection associated with disease severity and outcome. Science Immunology 07 Dec 2020: Vol. 5, Issue 54, eabe0240.

21. Quattrone F, Vabanesi M, Borghini A, De Vito G, Emdin M, Passino C. The value of hospital personnel serological screening in an integrated COVID-19 infection prevention and control strategy. Infect Control Hosp Epidemiol. 2020 May 15:1 –2.

22. Jalali Nadoushan M, Ahmadi S, Jalali Nadoushan P. Serology Testing for SARS-CoV-2: Benefits and Challenges. Iran J Pathol. 2020 Summer;15(3):154 –155.

23. Tahmina Shirin,Taufiqur Rahman Bhuiyan, Richelle C. Charles, Shaheena Amin,Imran Bhuiyan,Zannat Kawser et.al. Antibody responses after COVID-19 infection in patients who are mildly symptomatic or asymptomatic in Bangladesh. Int J Infect Dis. 2020 Dec; 101: 220–225.

24. Antonio La Marca, Martina Capuzzo,Tiziana Paglia,Laura Roli,Tommaso Trenti,and Scott M. Nelson.Testing for SARS-CoV-2 (COVID-19): a systematic review and clinical guide to molecular and serological in-vitro diagnostic assays. Reproductive biomedicine online, 41(3), 483–499.

25. Whitman, J. D., Hiatt, J., Mowery, C. T., Shy, B. R., Yu R., Yamamoto, et.al. Test performance evaluation of SARS-CoV-2 serological assays. medRxiv, 2020.04.25.20074856.

26. Gao Yong, Yuan Yi, Li Tuantuan, Wang Xiaowu, Li Xiuyong, Li Ang, Han Mingfeng. Evaluation of the auxiliary diagnosis value of antibodies assays for the detection of novel coronavirus (SARS-Cov-2), medRxiv 2020.03.26.20042044

27. Corine H. GeurtsvanKessel, Nisreen M.A. Okba, Zsofia Igloi, Carmen W.E. et.al. Towards the next phase: evaluation of serological assays for diagnostics and exposure assessment. medRxiv. 2020, 04.23.20077156

28. Li, Z., Yi, Y., Luo, X., Xiong, N., Liu, Y., Li, S, ET.AL Development and clinical application of a rapid IgM-IgG combined antibody test for SARS-CoV-2 infection diagnosis. Journal of medical virology, 92(9), 1518–1524.

29. Montesinos I, Gruson D, Kabamba B, Dahma H, Van den Wijngaert S, Reza S, Carbone V, et.al. Evaluation of two automated and three rapid lateral flow immunoassays for the detection of anti-SARS-CoV-2 antibodies. J Clin Virol. 2020 Jul; 128:104413

30. Barnaby Flower,1,2 Jonathan C Brown,1 Bryony Simmons,1 Maya Moshe,1 Rebecca Frise,1 Rebecca Penn et.al.Clinical and laboratory evaluation of SARS-CoV-2 lateral flow assays for use in a national COVID-19 seroprevalence survey. Thorax 2020;75:1082–1088.

31. Wu, J. L., Tseng, W. P., Lin, C. H., Lee, T. F et.al. Four point-of-care lateral flow immunoassays for diagnosis of COVID-19 and for assessing dynamics of antibody responses to SARS-CoV-2. The Journal of infection, 81(3), 435–442.

32. Mayara Lisboa Bastos, Gamuchirai Tavaziva, Syed Kunal Abidi, Jonathon R Campbell, Louis-Patrick Haraoui and James C Johnston. Diagnostic accuracy of serological tests for covid-19: systematic review and meta-analysis. BMJ 2020;370:m2516.

33. Anna Christine Nilssona, 1, Dorte Kinggaard Holma, Ulrik Stenz Justesenb, Thøger Gorm-Jensenb,Nc anna Skaarup Andersenb, Anne Øvrehus et.al. Comparison of six commercially available SARS-CoV-2 antibody assays—Choice of assay depends on intended use. nt J Infect Dis. 2021 Feb; 103:381–388.

34. Espejo, A. P., Akgun, Y., Al Mana, A. F., Tjendra, Y., Millan, N. C., Gomez - Fernandez, C., & Cray, C. Review of Current Advances in Serologic Testing for COVID-19. American journal of clinical pathology, 154(3), 293–304.

